# Effective connectivity of emotion and cognition under psilocybin

**DOI:** 10.1101/2022.09.06.22279626

**Authors:** Devon Stoliker, Leonardo Novelli, Franz X. Vollenweider, Gary F. Egan, Katrin H. Preller, Adeel Razi

## Abstract

Classic psychedelics alter sense of self and patterns of self-related thought. These changes are hypothesised to underlie their therapeutic efficacy across internalising pathologies such as addiction and depression. Using resting-state functional MRI images from a randomised, double blinded, placebo-controlled clinical trial of 24 healthy adults under 0.215mg/kg psilocybin, we investigated how psilocybin modulates the effective connectivity between resting state networks and the amygdala that are involved in the appraisal and regulation of emotion and association with clinical symptoms. The networks included the default mode network (DMN), salience network (SN) and central executive network (CEN). Psilocybin decreased top-down effective connectivity from the resting state networks to the amygdala and decreased effective connectivity within the DMN and SN, while the within CEN effective connectivity increased. Effective connectivity changes were also associated with altered emotion and meaning under psilocybin. Our findings identify changes to cognitive-emotional connectivity associated with the subjective effects of psilocybin and the attenuation of the amygdala signal as a potential biomarker of psilocybin’s therapeutic efficacy.

## Introduction

Psilocybin, the main active compound of so-called magic mushrooms, is a serotonergic psychedelic that has been used throughout history in religious and medical rituals to alter consciousness (Carod-Artal, 2015; Nichols, 2016). It belongs to a larger group of substances such as ayahuasca (DMT) and mescaline that share molecular binding kinetics to serotonergic 5-HT receptors distributed throughout the brain (Nichols, 2012). Agonism of the 5-HT2A receptor is primarily responsible for psychedelic subjective effects that can cause powerful changes to subjective experience (Vollenweider et al 1998; (Kometer et al., 2012) Kometer et al. 2013; Preller et al. 2017; Kraehenmann et al. 2017). Psychedelics can also produce feelings of subjective wellbeing (Hirschfeld & Schmidt, 2020) and profound changes in self-awareness and perception, including visual hallucinations and synaesthesia, and a sense of transcendence of time and space (Studerus, Gamma, & Vollenweider, 2010; Swanson, 2018; Wittmann et al., 2007; (Stoliker, Egan, Friston, & Razi, 2022). Altered (self-)perception and self-awareness are unique effects of serotonergic psychedelics and may involve an experience of ego dissolution, defined as a loss of self-boundaries and sense of unity between the self and the environment (Stoliker, Egan, Friston, & Razi, 2021; Stoliker, Egan, Friston, et al., 2022)Lebedev et al., 2015). Although complete ego dissolution is rare at the medium dose potency used in clinical trials, several studies have found that overall blissful ego dissolution (as measured by the total mystical score or oceanic self-boundlessness) is associated with better long-term outcomes in healthy subjects (Griffiths et al., 2011) and reduced symptoms of depression (Griffiths et al., 2016; Roseman, Nutt, & Carhart-Harris, 2018; Ross et al., 2016) and addiction (Bogenschutz et al., 2015; Garcia-Romeu, Griffiths, & Johnson, 2014).

Depression has been linked to a fundamental sense of disconnectedness (Northoff, 2007), which may be ameliorated by psychedelics. Studies have found reduced depression scores and improved wellness associated with enhanced the subjective sense of connectedness following an acute psychedelic experience (R. L. Carhart-Harris et al., 2018; Watts, Day, Krzanowski, Nutt, & Carhart-Harris, 2017; Watts et al., 2022). Research findings indicate a strong correlation between ratings of psychological insight one day after a session with psilocybin and decreases in depression four weeks later (Alan K. Davis et al., 2021). Reduced depression may also be mediated by increased cognitive and psychological flexibility after psychedelics (A. K. Davis, Barrett, & Griffiths, 2020; Doss et al., 2021). These findings suggest that the long-term benefits of psychedelics for mental health may result from modulation of the regulation of emotions and cognitive processing (Duerler, Vollenweider, & Preller, 2022; Franz X. Vollenweider & Preller, 2020). Moreover, the acute psychedelic experience appears to be crucial to these changes.

The amygdala is a bilateral subcortical region composed of many subnuclei that integrate cortical and thalamic sensory inputs (Babaev, Piletti Chatain, & Krueger-Burg, 2018). Its primary function is to respond to the emotional significance of stimuli and regulate arousal (Alexander et al., 2021; Bonnet et al., 2015). For example, presentations of positive or negative emotional stimuli, such as physical threats, elicit activation of the amygdala (Alexander et al., 2021; Garavan, Pendergrass, Ross, Stein, & Risinger, 2001; Hobin, Goosens, & Maren, 2003; Weymar & Schwabe, 2016). The amygdala also responds to cognitive-emotional significance. For example, when personal beliefs are threatened (Kaplan, Gimbel, & Harris, 2016). This demonstrates self-beliefs can condition and regulate amygdala emotional responses. Similarly, complex emotions such as shame are associated with the amygdala and can be diminished by its damage (Piretti et al., 2020). Emotional empathy is also associated with amygdala function (Geng et al., 2018; Hillis, 2014; Marsh et al., 2013), and has been reported to be modified by psilocybin (Pokorny, Preller, Kometer, Dziobek, & Vollenweider, 2017; Preller et al., 2015).

Further associations between emotional and behaviour regulation and the amygdala are demonstrated by damage to the amygdala which affects decision-making (Gupta, Koscik, Bechara, & Tranel, 2011) and memory (Adolphs, Tranel, & Denburg, 2000). Amygdala hyperactivation is also a marker of stress and anxiety, which are implicated across internalising disorders for which psychedelics demonstrate efficacy (Nutt & Carhart-Harris, 2020); Koob & Volkow, 2010; Luo, Xue, Shen, & Lu, 2013; Menon, 2011). Emotional regulation depends on top-down and bottom-up modulation of the amygdala (McRae, Misra, Prasad, Pereira, & Gross, 2012; Ochsner et al., 2009) and appears to be altered by psychedelics. For example, healthy adults administered psilocybin show reduced effective connectivity from the amygdala to the primary visual cortex coincident with reduced threat processing (Kraehenmann et al., 2015) (Kraehenmann et al., 2016). The semisynthetic psychedelic drug lysergic diethylamide acid (LSD) has also been shown to dampen the amygdala response to fearful stimuli (Mueller et al., 2017; Barrett, Doss, Sepeda, Pekar, & Griffiths, 2020). Furthermore, decreased amygdala activity has been correlated with positive mood under psilocybin lasting up to one month (Kraehenmann et al., 2015; Barrett et al., 2020). However, subjects with treatment resistant depression imaged one day post-treatment show increased amygdala responses to stimuli with psilocybin as well as improvements in depressive symptoms (Roseman, Demetriou, Wall, Nutt, & Carhart-Harris, 2018; Barrett et al., 2020). Together, these results suggest a change in amygdala connectivity under the acute influence of psychedelics may be a biological mechanism underwriting the beneficial, mood-enhancing effects of psilocybin.

The amygdala is integrated with cognition through top-down projections from midline cortical regions (Etkin et al., 2011; Kraehenmann et al., 2016; Robinson et al., 2013). These projections are thought to underlie the perception and intensity of positive and negative emotions (Bonnet et al., 2015) and abnormal connectivity of this circuitry underlies psychopathological conditions such as anxiety disorders (Menon, 2011). Amygdala connectivity changes that involve frontal cortical regions have been found in several studies suggesting psilocybin modulates the top-down control of limbic regions involved in emotional processing (Bernasconi et al., 2014; Mertens et al., 2020; Mueller et al., 2017; Barrett et al., 2020). Changes to the amygdala and cortical regions are sustained well beyond the half-life of psilocybin and may last up to one month post administration (Barrett, Doss, Sepeda, Pekar, & Griffiths, 2020), demonstrating the lasting neural changes in association with increased positive affect.

Emotion is a flexible, dynamic and functionally integrated with cognitive regions of the brain (Lindquist, Satpute, Wager, Weber, & Barrett, 2016). The function of cortical regions linked to amygdala are related to the sense of self, self-referential cognition, cognitive appraisal, and emotional regulation. For example, limbic-cortical dysregulation has been reported to impact mood regulation (Zeng et al., 2012). The changes to cortical regions under the acute effects of psychedelics may modulate subcortical regions that disrupt affective responses that may potentially support therapeutic responses.

Here we investigate to what extent psilocybin modulates the relationship of cognitive-emotional networks interactions that may be relevant for therapeutic outcomes. We investigated amygdala connectivity to regions that form frontal and anterior nodes of distinct large-scale resting-state networks (RSNs). RSNs are determined by temporally correlated activation across spatially distributed brain regions and serve essential functions in cognition (Fox et al., 2005; Raichle, 2015). The RSN regions of particular relevance to this study are abundant in 5-HT2A receptors (Saulin, Savli, & Lanzenberger, 2012; R. L. Carhart-Harris & Friston, 2019) and include the default mode network (DMN), central executive network (CEN) and the salience network (SN).

The DMN integrates internal and external information to perform self-referential mental activity involved in forming self-beliefs (Amey, Leitner, Liu, & Forbes, 2022; Tompson, Chua, & Kitayama, 2016) and is composed of the medial prefrontal cortex (mPFC) and posterior cingulate cortex (PCC) central hub regions. Increased DMN connectivity with the amygdala is observed in depressed subjects (Sheline Yvette et al., 2009) and has been associated with symptoms such as rumination (Cooney, Joormann, Eugène, Dennis, & Gotlib, 2010). In contrast, decreased coupling of the central hub regions of the DMN was reported in association with the reduced mind wandering of advanced meditation practitioners (Judson A. Brewer et al., 2011). Similar decoupling was found acutely (Carhart -Harris et al 2012) and two days after psilocybin (R. L.; Smigielski, Scheidegger, Kometer, & Vollenweider, 2019). We hypothesise reduced interactions between the amygdala and DMN and reduced within-DMN connectivity may underpin an altered sense of self and self-related emotion under psychedelic subjective effects and these subjective effects can be measured using the blissful state (emotion) and changed meaning of percepts subscales of the 5D-ASC (Studerus, Gamma, & Vollenweider, 2010). Connectivity changes, particularly those associated with subjective changes, may serve as a biomarker of connectivity that may have therapeutic properties.

The central executive network (CEN) is a RSN involved in thinking, planning, and controlling attention (Andrews-Hanna et al., 2014; Menon, 2011). Abnormal CEN function is associated with mental health in various cognitive disorders. For example, reduced CEN activity has been associated with chronic schizophrenia and depression (Leptourgos et al., 2020; Menon, 2011) and increased CEN-amygdala connectivity was observed in association with catastrophising of chronic pain (Jiang et al., 2016). The central hub regions of the CEN are the dorsolateral prefrontal cortex (dorsolateral PFC) and the lateral posterior parietal cortex (lateral PPC). The dorsolateral PFC serves a primary role in regulating emotion (Underwood et al., 2021) and was associated with worry in healthy adults and individuals with generalised anxiety (Paulesu et al., 2010; Servaas et al., 2014; Jamieson, Harrison, Razi, & Davey, 2022). Under psychedelics, a number of brain mapping studies suggest psychedelics increase the global connectivity and flexibility of CEN connectivity that may be associated with subjective emotional changes (Luppi et al., 2021; Tagliazucchi et al., 2016); Lord et al., 2019).

The salience network (SN) is a cognitive RSN involved in the detection of salient features in the environment and disengages task-irrelevant activity associated with the DMN, for example during daydreaming. The SN’s cardinal regions are the dorsal anterior cingulate cortex (dorsal ACC) and anterior insula (AI). Expanded network compositions of the SN also include the amygdala which may be important for the initial emotional appraisal of salient stimuli. Overactive salience attributed to hyperactivity of the AI or amygdala stimuli is a feature of various forms of psychopathology (Menon, 2011) and the AI is important to integrate affective and cognitive signals that influence interoceptive responses related to psychosomatic symptoms and anxiety and depression (Paulus, 2006; Terasawa, Shibata, Moriguchi, & Umeda, 2013; Wiebking et al., 2015). The dorsal ACC region of the SN is activated in response to social pain (Rotge et al., 2015) that has been associated with changes in self-processing (Smigielski et al. 2019; Smigielski et al., 2020) as well as reduced sense of social exclusion under psilocybin (Preller et al., 2016; Preller et al., 2015). Observations under psilocybin have also shown reduced SN integrity (Lebedev et al., 2015) and changes to effective connectivity from the SN to the DMN under LSD (Stoliker, Novelli, et al., 2021). Psychedelics may alter salience functions that select the pre-reflective channels of attention and modulate amygdala connectivity involved in emotional attention and appraisal (Kapur, 2003; Stoliker, Egan, et al., 2021).

## Results

### Network effective connectivity change with the amygdala under psilocybin

Participants underwent resting-state fMRI scans 70 minutes post oral administration of psilocybin. Effective connectivity between the amygdala and the DMN, SN and CEN and brain-behaviour analysis was performed at the group level using the framework of parametric empirical Bayes (PEB, see supplementary material for details). The reported results for effective connectivity and brain-behaviour analysis used a threshold of posterior probability > 0.99 for inference with very strong evidence. Those connections and associations not reported did not exceed this threshold. For more statistical details, see Supplementary Table 1.

#### i) Change of DMN effective connectivity to the amygdala under psilocybin

Group level effective connectivity strength between regions of the DMN and amygdala showed decreased self-inhibition of the PCC (effect size = −0.21) and decreased effective connectivity from the PCC with the left amygdala (effect size = −0.33 Hz) compared to placebo.

#### ii) Change of CEN effective connectivity to the amygdala under psilocybin

Several changes in effective connectivity were found, whose interplay is captured schematically in Fig. 2. Specifically, group level effective connectivity strength between regions of the CEN and amygdala showed increased self-inhibition of the right dorsolateral PFC (effect size = +0.17), left dorsolateral PFC (effect size = +0.19) and decreased self-inhibition of the left lateral PPC (effect size = −0.2), compared to placebo. Effective connectivity from the left lateral PPC to the left amygdala decreased (effect size = −0.22 Hz) and increased from right lateral PPC to the right dorsolateral PFC (effect size = +0.2 Hz) and decreased to the right amygdala (effect size = −0.39 Hz). Left dorsolateral PFC effective connectivity to the right amygdala decreased (effect size = −0.16 Hz) and increased to the left lateral PPC (effect size = +0.23 Hz), while right dorsolateral PFC increased to the left dorsolateral PFC (effect size = +0.15 Hz) and the right amygdala increased to the left lateral PPC (effect size = +0.28 Hz), compared to placebo.

**Fig 1.**
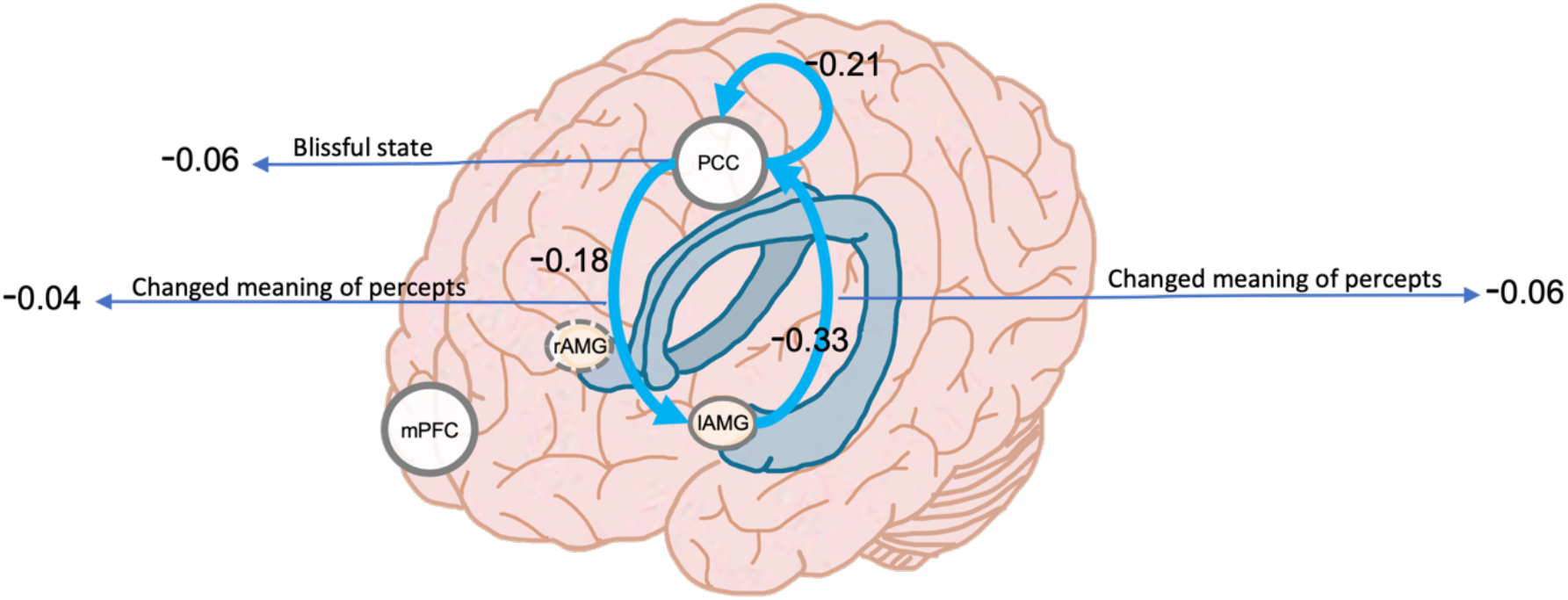
Default mode network effective connectivity change under psilocybin 70 minutes post-administration. Connections show changes in effective connectivity compared to placebo. Values display effect sizes (posterior expectations) of connections in Hz (except the inhibitory self-connections which are log-scaled). Values linked to subjective effects represent their associations with effective connectivity and represent normalized beta (β) coefficients. All results are for posterior probability > 0.99 (amounting to very strong evidence).

**Fig 2.**
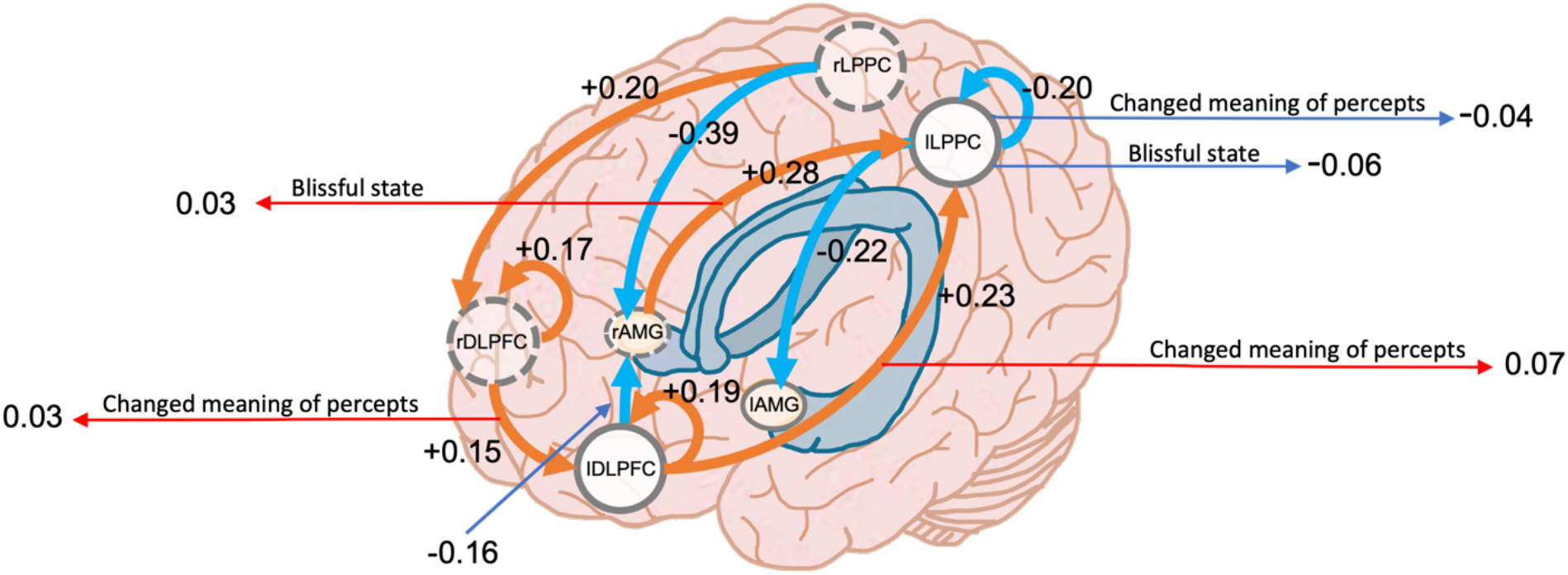
Central executive network effective connectivity change under psilocybin 70 minutes post-administration. Values display effect sizes (posterior expectations) of connections in Hz (except the inhibitory self-connections which are log-scaled). Values linked to subjective effects represent their associations with effective connectivity and represent normalized β coefficients. All results are for posterior probability > 0.99.

#### iii) Change of SN effective connectivity to the amygdala under psilocybin

Group-level effective connectivity showed analysis showed increased self-inhibition of the right AI (effect size = +0.23) and bilateral amygdala (effect size = +0.16 (right) and +0.31 (left)) and decreased effective connectivity from the dorsal ACC to right amygdala (effect size = −0.19 Hz) and from left AI to the right amygdala (effect size = −0.24 Hz) compared to placebo (Fig 3).

**Fig 3.**
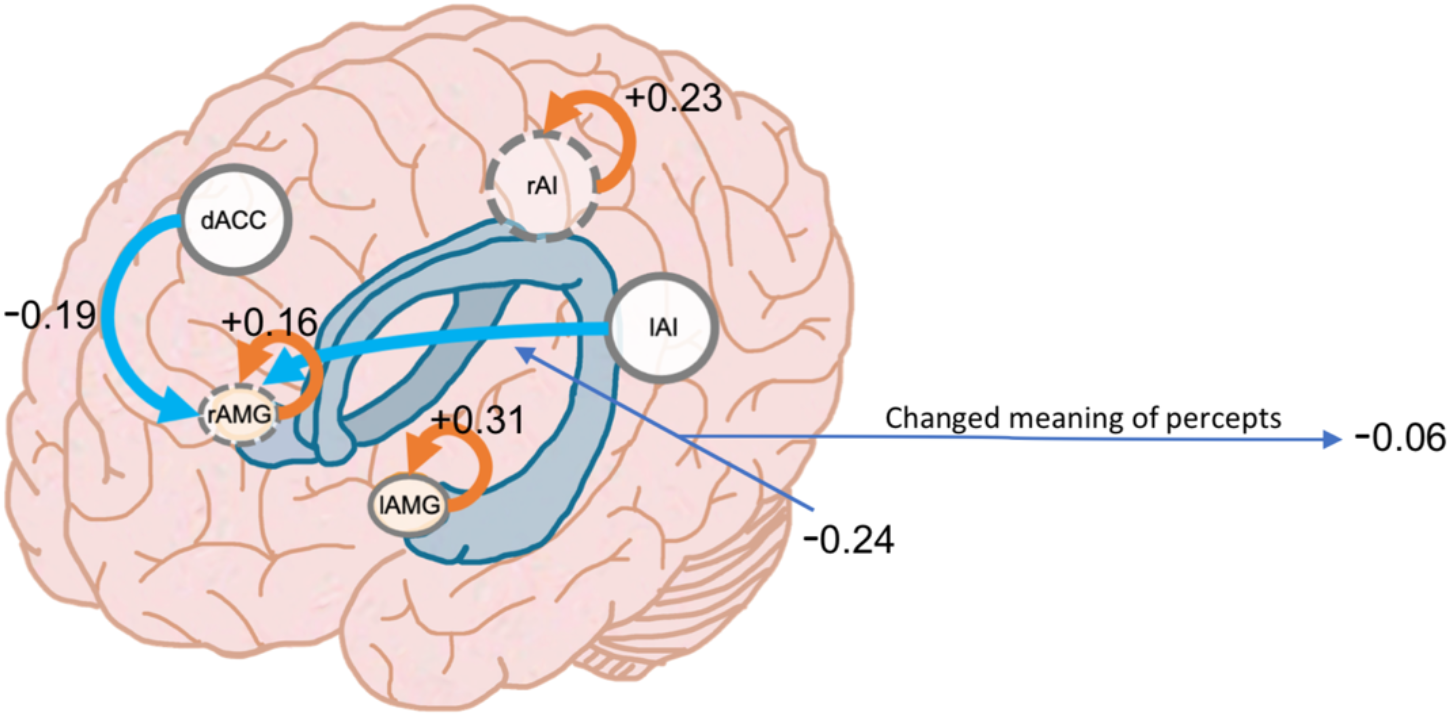
Salience network effective connectivity change under psilocybin 70 minutes post-administration. Connections show changes in effective connectivity compared to placebo. Values display effect sizes (posterior expectations) of connections in Hz (except the inhibitory self-connections which are log-scaled). Values linked to subjective effects represent their associations with effective connectivity and represent normalized β coefficients. All results are for posterior probability > 0.99.

### Brain-behaviour associations

#### i) Behavioural associations to DMN-amygdala effective connectivity change under psilocybin

The brain-behaviour analysis showed negative association between self-connectivity of PCC and blissful state (β coefficient = - 0.06), negative association between effective connectivity from PCC to left amygdala and changed meaning of percept (β coefficient = - 0.04), and negative association between effectivity from left amygdala to PCC and changed meaning of percept (β coefficient = - 0.06). See Fig 1.

#### ii) Behavioural associations to CEN-amygdala effective connectivity change under psilocybin

The brain-behaviour analysis showed positive association between effective connectivity from left lateral PPC to right amygdala and blissful state (β coefficient = + 0.03), positive association between effective connectivity from right DLPFC to left DLPFC and changed meaning of percept (β coefficient = + 0.03), negative association between self-connectivity of left lateral PPC to changed meaning of percept (β coefficient = - 0.04) ad blissful state (β coefficient = - 0.06), and positive association between effective connectivity from left DLPFC to left lateral PPC and changed meaning of percept (β coefficient = + 0.07). See Fig 2.

#### iii) Behavioural associations to SN-amygdala effective connectivity change under psilocybin

The brain-behaviour analysis showed negative association between effective connectivity from left AI to right amygdala and changed meaning of percept (β coefficient = - 0.06). See Fig 3.

## Discussion

To investigate the influence of psilocybin on the interaction between emotion and cognition, we measured connectivity changes between the amygdala and selected RSNs. These networks serve functions in selfhood and cognition and are composed of regions linked to the regulation and experience of emotion. Our results demonstrate increased inhibitory effective connectivity from cortical regions to the amygdala under psilocybin. For example, we showed this change in connectivity from the PCC, left dorsolateral PFC, bilateral lateral PPC, dorsal ACC, and left AI. These results suggest cognitive resources do not recruit the amygdala under psilocybin, and this modulation of top-down connectivity was related to subjective effects of psilocybin.

Examination of the DMN-amygdala connectivity changes under psychedelics did not find effective connectivity change involving the mPFC, which has been frequently cited in association with emotion, such as emotional valence (Willinger et al., 2019; Underwood, Tolmeijer, Wibroe, Peters, & Mason, 2021). However, examination of the PCC, which orients information towards the self (Andrews-Hanna, Smallwood, & Spreng, 2014; J. A. Brewer, Garrison, & Whitfield-Gabrieli, 2013; Qin & Northoff, 2011), and amygdala revealed connectivity changes and behavioural associations under psilocybin. PCC and amygdala effective connectivity change was bidirectional and accompanied by reduced PCC self-inhibition, which indicates increased synaptic gain or its sensitivity to inputs. The increased PCC synaptic gain was associated with the subjective experience of blissful state, and PCC bidirectional inhibition with the left amygdala was associated with changed meaning of percepts. Our findings suggest inhibition between the amygdala and PCC may be an important connectivity change underlying altered meaning (i.e., everyday things gained symbolic, strange, or emotional meaning), that, in turn, may contribute to the change in self-awareness. Our findings also indicate PCC decreased inhibition facilitates blissful feelings. These changes may reflect reduced top-down cognitive control associated with loss of self-referential processing that is experienced as a dissolution of self-boundaries and align with the proposed role of the PCC in self-referential processing (Brewer, Garrison, & Whitfield-Gabrieli, 2013; Qin & Northoff, 2011) as well as previous evidence of PPC change associated to the dissolution of self-boundaries under psychedelics (Muthukumaraswamy et al., 2013). For example, previous findings found PCC self-inhibition under LSD, and the PCC’s association with diminished self-representation in social interaction, feelings of self and self-other processing under psychedelics (Preller et al., 2017; Preller et al., 2019; Preller et al., 2018; Stoliker, Novelli, et al., 2022).

Left dorsolateral PFC and left lateral PPC inhibition to the amygdala was also observed and is situated within the CEN. Connectivity in this network is believed to underlie the evaluative aspect of thought, with the dorsolateral PFC playing a central role in emotional regulation. However, the observed increased top-down inhibition of this connectivity to the amygdala was not associated with subjective effects assessed with the 5D-ASC scale. Instead, the pattern of activity in this network is open to interpretation. One characterisation of the observed activity may be its resemblance to hyper-frontality, which is associated with worry and anxiety (Park, Wood, Bondi, Del Arco, & Moghaddam, 2016), and acute psychosis (F. X. Vollenweider et al., 1997). There is also evidence that CEN connectivity is increased, and the amygdala connectivity is decreased, during cognitive control strategies (Herwig et al., 2007; Herwig, Kaffenberger, Jäncke, & Brühl, 2010). However, links between CEN activity and behaviour are suggested to be task and context-dependent (Borders, 2020) and subjects in this study reported nominal anxiety, see Supplementary Material. An alternative interpretation is that the observed connectivity pattern between regions of the CEN constitutes an increase in evaluative thought processes and may be linked to re-appraisal. For example, increased CEN and decreased amygdala activity of healthy adults was observed to increase during emotion-introspection, while increased CEN and increased amygdala activity was associated with self-reflection (Herwig et al., 2010). This literature may help explain the activation differences in the amygdala observed between healthy adults under psychedelics and clinical populations in the days after psychedelics. Moreover, increased CEN and decreased amygdala activity align our findings with connectivity underlying emotion-introspection. Although the introspection of healthy adults in this imaging study was not assumed, psilocybin experiences are commonly related to emotional introspection. Further evidence demonstrating the therapeutic capacity of increased intrinsic CEN connectivity is presented in association with resilience (Miller Gregory et al., 2018). Moreover, positive self-appraisal has been associated with increased CEN and amygdala connectivity (Brühl, Rufer, Kaffenberger, Baur, & Herwig, 2014), which may relate to self-reflection in the days after psilocybin administration. This evidence indicates the increased CEN connectivity and CEN top-down inhibition to the amygdala as a connectivity change that relates to the therapeutic potential of psilocybin for the revision of beliefs and it warrants further investigation.

Although we expect re-evaluation of thought under psychedelics to be influenced by the context (i.e., the mindset of participants and environmental setting), measured subjective effects associated with CEN connectivity changes are detectable in the imaging environment. For example, increased connectivity from the left dorsolateral PFC to the left lateral PPC and connectivity between the bilateral dorsolateral PFC was associated with changed meaning of percepts. Moreover, connectivity from the left dorsolateral PFC and right AMG to the left lateral PPC and decreased left lateral PPC self-inhibition were associated with both blissful state and the changed meaning of percepts. The lateral PPC inhibition to the amygdala may suggest its decoupling from the amygdala limits the sensory and interoceptive information to the amygdala, while the left amygdala synaptic gain increased. These results demonstrate nuanced modulation of the CEN-amygdala connectivity that align with previous reports of functional desegregation and expanded access to wider dynamics of brain states made available under psychedelics (Atasoy et al., 2017; Barnett, Muthukumaraswamy, Carhart-Harris, & Seth, 2019; R. L. Carhart-Harris & Nutt, 2017; A. K. Davis et al., 2020; Matias, Lottem, Dugué, & Mainen, 2017; Stoliker, Egan, & Razi, 2022; Tagliazucchi, Carhart-Harris, Leech, Nutt, & Chialvo, 2014; Watts & Luoma, 2020). These connectivity changes may be important to psilocybin’s therapeutic potential that can be targeted to facilitate cognitive-emotional changes.

The ACC and AI serve essential functions in salience detection, and the integration of these regions with the amygdala may underwrite pathologically enhanced salience detection (Menon, 2011). These regions’ inhibition of the right amygdala suggests their role in salience detection is diminished under psilocybin, which may be therapeutic for internalising disorders. Notably, previous literature found that psilocybin acutely reduced mainly the response in the right amygdala to negative stimuli, which may reflect “decentering” from one’s emotion and thought that may facilitate reappraisal (Malinowski, 2013). However, inhibited salience detection also aligns with positive symptoms in schizophrenia (i.e., hallucinations and delusions) (Menon, 2011). At higher doses, psychedelics can induce a psychotic-like state in some individuals (F. X. Vollenweider, Vollenweider-Scherpenhuyzen, Babler, Vogel, & Hell, 1998), that may be related to the effect of serotonergic psychedelics on salience detection (Preller et al., 2017). However, psychotic-like behavioural outcomes rarely occur in a controlled clinical setting (Johnson, Richards, & Griffiths, 2008; Studerus, Gamma, Kometer, & Vollenweider, 2012; Studerus et al., 2010).

Previous research shows that increased functional connectivity between the AI and amygdala is associated with relapse of addiction in preclinical studies (Venniro et al., 2017) and behavioural habituation in healthy adults (Denny et al., 2014). Increased effect connectivity of this link has been associated with depression in clinical populations (Kandilarova, Stoyanov, Kostianev, & Specht, 2018). The AI inhibition to the amygdala was associated with changed meaning of percepts that suggests it may be associated with therapeutic behavioural change. Previous task-based research shows reduced dorsal ACC activity in association with blunted response to social exclusion under psilocybin (Preller et al., 2015). Although our findings did not associate subjective effects with this connection, dorsal ACC-related inhibition aligns with the inhibition of social pain (Preller et al., 2016; Preller et al., 2015) and previous findings of this circuit implicate top-down inhibition in the resolution of emotional conflict (Etkin, Egner, Peraza, Kandel, & Hirsch, 2006). DCM also estimated increased self-inhibition in the bilateral amygdala and rAI, suggesting reduced synaptic gain of the regions. LSD produced similar results that demonstrated hypo-connectivity of the right insula (Preller et al., 2018). However, the changes we observed were not associated with the measured psychedelic subjective effects.

Taken together, our results suggest that psilocybin attenuates the recruitment of the amygdala. This connectivity contributes to the altered meaning of percepts and subjective emotion. Diminished amygdala activation has been related to emotional regulation (Herwig et al., 2019) and may alter the patterns of connectivity that condition emotional, cognitive and self-reflective responses. Psilocybin induced change to the RSNs that we observed generally aligns with previous literature that shows the increased flexibility and transition of network states under psychedelics, as well as nimble and flexible brain networks are instrumental to positive emotion and affect (Alexander et al., 2021). Therefore, the RSNs reduced connectivity to the amygdala and altered within-RSN effective connectivity that we observed may be important aspects of the subjective effects and emotional-cognitive connectivity changes that therapeutic applications capitalise upon to encourage revision of beliefs.

Our endeavour to investigate interactions between emotion and cognition in a healthy population under psilocybin has several limitations. For example, our analysis would benefit from MRI scans taken before and after the administration of psilocybin. Exploring links between emotional and associative connectivity also involves deciding network configuration and regions for analysis. Regions such as the ventral striatum, precuneus, and parahippocampus and further subregions of the PFC and ACC may also be valuable for examining the links between emotion and cognition (Underwood et al., 2021). Furthermore, each region selected as part of the network may be determined differently depending on the method of identification of the region. Further standardisation of this process is complex for anatomical reasons and remains under debate. The processing pipeline, participant sample and dose of psilocybin can also strongly impact the reliability of results. Participants were given lower doses than currently used in clinical studies of depression and substance dependence disorders. Therefore, connectivity dynamics may be dissimilar at a higher dose. Data sets at higher doses may provide improved inference and more nuanced characterisation of the neural dynamics of interest. We also overlooked the influence of numerous neurotransmitter systems involved in emotion that may interact with altered serotonin transmission. These interactions may provide a novel line of inquiry to understand psychedelic therapeutic effects.

Other limitations in the methods and analysis include the use of global signal regression and the limited temporal resolution. Temporal resolution is likely to be significant in the study of emotion. For example, the time course of emotion may present distinct contributions of brain regions involved in valence and arousal (Alexander et al., 2021). Modalities with temporal specificity capable of resolving insight and emotional change, such as EEG or MEG, which can also be analysed using DCM, are needed in future investigations. Furthermore, studies using task-based fMRI paradigms can induce activations in specific emotion and cognition related brain regions (Brühl et al., 2014; Herwig et al., 2007; Herwig et al., 2010).

The generalisability of our results to clinical populations is limited by the use of a small healthy adult sample and the congruence between our results and previous findings. Previous research indicates psychedelics may globally increase bottom-up connectivity (R. Carhart-Harris, Kaelen, & Nutt, 2014; R. L. Carhart-Harris & Friston, 2019; Swanson, 2018). For example, evidence of thalamic connectivity to the cortex supports this view (Müller, Dolder, Schmidt, Liechti, & Borgwardt, 2018; Preller et al., 2019). However, decoupling between limbic regions and the cortex has also been identified (Robin L. Carhart-Harris et al., 2016; Lebedev et al., 2015). Our findings that identified increased bottom-up connectivity from the right amygdala to the left lateral PPC align with the latter studies. As psychedelic research advances, we suggest examination of multiple limbic regions and their subnuclei connectivity to the cortex. This may reveal more precise and nuanced dynamics between the subcortex and cortex underlie previously theorised global patterns. For example, recent investigations have demonstrated spatial specificity of entropic activity and thalamic connectivity associated with psychedelic subjective effects (Gaddis et al., 2022). Along these lines, the amygdala is composed of many subnuclei. Its small size, anatomical variability and position deep within the brain presents challenges for noise-to-signal ratio and spatial location in group fMRI analysis, which future imaging and research methods may overcome. Finally, increased excitation among the CEN effective connectivity contrasts with previous analyses, which found a general reduction in associative functional connectivity (Preller et al., 2020). While the global reduction of associative connectivity may help explain experiences of ego dissolution, psychedelic experiences also contain phenomenological richness (Millière, Carhart-Harris, Roseman, Trautwein, & Berkovich-Ohana, 2018). Future research of CEN regions may wish to address the reliability of this effective connectivity and examine links between the CEN, subjective experience, and therapeutic outcomes. Such work may benefit from finer grained emotional and cognitive rating scales.

In sum, we demonstrated altered within-RSN effective connectivity and reduced top-down RSN-amygdala effective connectivity associated with emotional and cognitive behavioural changes under psilocybin. The functional significance of these changes to perceptual processes and psychopathology may be reduced cognitive control and increased flexibility of these networks and the attenuated modulation of the amygdala under the acute effects of psilocybin. These changes may enable a mental workspace for therapeutic emotional and cognitive change. We suggest RSN attenuation of the amygdala as a potential biomarker for future clinical research investigating the neural correlates of psychedelic therapeutic efficacy. Future work may also build upon our effective connectivity findings to understand how connectivity underlying emotion and cognition interact with the therapeutic context to effect patterns of thought and behaviour. These lines of enquiry can help discern the imperative of the psychedelic subjective experience to therapeutic outcomes and bridge gaps in our understanding of the relationship between brain connectivity and behaviour.

## Methods

### Participants

The data analysed in this paper were collected as part of a previous study (registered at ClinicalTrials.gov (NCT03736980)), which is reported in (Preller et al., 2020) and was approved by the Cantonal Ethics Committee of Zurich. Twenty-four subjects (12 males and 11 females; mean age = 26.3 y; range = 20–40 y; 1 subject did not complete the study) were recruited through advertisements at universities in Zurich, Switzerland. All participants were deemed healthy after screening for medical history, physical examination, blood analysis, and electrocardiography. The Mini-International Neuropsychiatric Interview (MINI-SCID) (Sheehan et al., 1998), the DSM-IV fourth edition self-rating questionnaire for Axis-II personality disorders (SCID-II) (Fydrich, Renneberg, Schmitz, & Wittchen, 1997), and the Hopkins Symptom Checklist (SCL-90-R) (Franke, 2002) were used to exclude subjects with present or previous psychiatric disorders or a history of major psychiatric disorders in first-degree relatives. Participants were asked to abstain from prescription and illicit drug use two weeks prior to first testing and throughout the duration of the study, and abstain from alcohol use 24 hours prior to testing days. Urine tests and self-report questionnaires were used to verify the absence of drug and alcohol use. Urine tests were also used to exclude pregnancy. Further exclusion criteria included left-handedness, poor knowledge of the German language, cardiovascular disease, history of head injury or neurological disorder, history of alcohol or illicit drug dependence, MRI exclusion criteria, including claustrophobia, and previous significant adverse reactions to a hallucinogenic drug. All participants provided written informed consent statements in accordance with the declaration of Helsinki before participation in the study. Subjects received written and oral descriptions of the study procedures, as well as details regarding the effects and possible risks of drug treatment.

### Design

A double blind, randomised, placebo-controlled, cross-over study was performed. Testing days occurred two weeks apart and participants were orally administered either psilocybin (0.2mg/kg orally) or placebo (179 mg mannitol and colloidal silicon dioxide (Aerosil) 1 mg. Resting state scans (10 minutes each) were taken 20, 40 and 70 minutes following administration. However, only scans at 70 minutes during the peak effects were used in our analysis. Participants were asked to not engage in repetitive thoughts such as counting and instructed to close their eyes during the resting state scan. Subjective experience was assessed using the retrospective 5-Dimensions Altered States of Consciousness (5D-ASC) questionnaire. Participants completed the 5D-ASC 360 min following psilocybin administration, see Supplementary.

### MRI Data Acquisition and Preprocessing

MRI data were acquired on a Philips Achieva 3T whole-body scanner. A 32-channel receive head coil and MultiTransmit parallel radio frequency transmission was used. Images were acquired using a whole-brain gradient-echo planar imaging (EPI) sequence (repetition time, 2,430 ms; echo time, 27 ms; slice thickness, 3 mm; 45 axial slices; no slice gap; field of view, 240 × 240 mm^2^; in-plane resolution, 3 × 3 mm^2^; sensitivity-encoding reduction factor, 2.0). 265 volumes were acquired per resting state scan resulting in a scan duration of 10 mins. Additionally, two high-resolution anatomical images (voxel size, 0.7 × 0.7 × 0.7 mm^3^) were acquired using T1-weighted and T2-weighted sequences using 3D magnetization prepared rapid-acquisition gradient echo sequence (MP-RAGE) and a turbo spin-echo sequence, respectively. See Supplementary for more details. The acquired images were analysed using SPM12 (https://www.fil.ion.ucl.ac.uk).

The pre-processing steps of the images consisted of slice-timing correction, realignment, spatial normalization to the standard EPI template of the Montreal Neurological Institute (MNI), and spatial smoothing using a Gaussian kernel of 6-mm full-width at half maximum. Head motion was investigated for any excessive movement. Three subjects were excluded due to head motion and one subject did not complete the scan at 70 minutes.

### Independent RSN-AMG DCM models

To reduce model complexity and focus the interpretation of the results on the connections of interest our analysis investigated independent models of amygdala effective connectivity with the DMN, CEN and SN.

### Extraction of region coordinates across subjects

Neurosynth, a large-scale automated synthesis of functional neuroimaging data, was used to identify coordinates composing regions of interest (Yarkoni, Poldrack, Nichols, Van Essen, & Wager, 2011). More details can be found in Supplementary Material.

Networks were composed of cardinal regions constituting core parts of the DMN (Andrews-Hanna, Reidler, Sepulcre, Poulin, & Buckner, 2010; Dixon et al., 2017), SN and CEN. The DMN and SN followed the selection of regions in a related investigation by Zhou and colleagues (Zhou et al., 2018) and all networks were included for their role in consciousness and link to emotion (Alexander et al., 2021; Borders, 2020; Underwood, Tolmeijer, Wibroe, Peters, & Mason, 2021). Associations between peak coordinates and cardinal nodes of network regions of interest (ROI) were determined by expert visual inspection. The MNI coordinates of the selected ROIs are listed in Table 1.

**Table 1.** Coordinates of regions of interest. The DMN comprised of the posterior cingulate cortex (PCC) and medial prefrontal cortex (mPFC); the SN comprised of the dorsal anterior cingulate cortex (dorsal ACC), left and right anterior insula (lAI/rAI); and the CEN comprised the left and right dorsal lateral prefrontal cortex (ldPFC/rd PFC) and the left and right lateral posterior parietal lobule (lIPPC/rlPPC).

| <u>Region</u> | <u>MNI coordinates</u> |  |  | <u>Network/System</u> |
| --- | --- | --- | --- | --- |
|  | x | y | z |  |
| PCC | 0 | -50 | 28 | DMN |
| mPFC | 0 | 54 | 20 | DMN |
| dACC | 2 | 28 | 22 | SN |
| lAI | -34 | 19 | 0 | SN |
| rAI | 38 | 18 | 0 | SN |
| ldPFC | -46 | 30 | 32 | CEN |
| rdPFC | 42 | 37 | 31 | CEN |
| lIPPC | -22 | -62 | 62 | CEN |
| rl PPC | 19 | -64 | 55 | CEN |
| lAMG | -21 | -5 | -17 | Limbic |
| rAMG | 23 | -3 | -19 | Limbic |

A generalized linear model (GLM) was used to regress 6 head motion parameters (3 translation and 3 rotational), white matter and cerebrospinal fluid signals from preprocessed data. One subject was excluded as no activation was found in one or more regions of interest. We also used global signal regression in our pre-processing pipeline in line with our previous work (Stoliker, Novelli, et al., 2021). The time series for each ROI was computed as the first principal component of the voxel activity within a 6 mm radius sphere centred on the ROI coordinates (as listed in Table 1).

### Specification and Inversion of DCM

Three independent fully-connected DCM models were specified for each network and the amygdala using the eleven ROIs defined in Table 4, without any exogenous inputs. The DCM for each subject was then inverted using spectral DCM (Karl J. Friston, Kahan, Biswal, & Razi, 2014; Razi, Kahan, Rees, & Friston, 2015) to infer the effective connectivity that best explains the observed cross-spectral density for each subject. This procedure was repeated for each testing condition. The DCM fit to the data using cross spectral density averaged 90.9% for placebo conditions and 89.8% for psilocybin conditions.

### Second Level Analysis Using Parametric Empirical Bayes

The effective connectivity inferred by spectral DCM for each subject was taken to the second (group) level to test hypotheses concerning between-group effects. A general linear model (GLM) was employed to decompose individual differences in effective connectivity into hypothesised group-average connection strengths together with unexplained noise. Hypotheses on the group-level parameters are tested within the parametric empirical Bayes (PEB) framework (K. J. Friston et al., 2016), where both the expected values and the covariance of the parameters were considered. That is, precise parameter estimates influence the group-level result more strongly than uncertain estimates, which are down-weighted. Bayesian model reduction (BMR) is used as an efficient form of Bayesian model selection (K. J. Friston et al., 2016). Please see supplementary material for further technical details about PEB.

## Data Availability

All data produced in the present study are available upon reasonable request to the authors.

## Author contributions

Conceptualization: DS, AR, FXV

Methodology: DS, LN, AR

Investigation: DS, LN, AR

Visualization: DS, LN, AR

Supervision: AR

Writing—original draft: DS

Writing—review & editing: DS, LN, AR, GE, KP, FXV

