## Supplementary Information for "Effective connectivity of emotion and cognition under psilocybin"

### Spectral Dynamic Causal Modelling

Dynamic causal modelling (DCM) is a Bayesian framework that infers the directed connectivity among the neuronal systems – referred to as effective connectivity. . Mathematically, the stochastic state-space generative model consists of two equations. First is the neuronal state differential equation, namely

$$\dot{x}(t) = f(x(t), u(t), \theta) + v(t), \quad (1)$$

and second is the observation equation, which is a static nonlinear mapping from the hidden physiological states in (1) to the observed BOLD activity and is written as:

$$y(t) = h(x(t), \varphi) + e(t), \quad (2)$$

where $\dot{x}(t)$ is the rate of change of the neuronal states $x(t)$, $\theta$ are the unknown effective connectivity parameters, and $v(t)$ is the state noise, i.e. a stochastic process that models the random neuronal fluctuations that drive the resting state activity. The term $u(t)$ represents any exogenous (or experimental) inputs that drive the hidden states – that are usually absent in resting state designs (Karl J. Friston, Kahan, Biswal, & Razi, 2014). In the observation equation, $\varphi$ are the unknown parameters of the haemodynamic observation function $h$ and $e(t)$ is the measurement (observation) noise.

This study employed spectral DCM for resting state fMRI, which simplifies the generative model by replacing the original timeseries with their second-order statistics (i.e., their cross spectra, which are the transformed cross-covariance terms in the frequency domain). Note that the cross-spectra are time invariant and their estimation is easier than that of the time-varying neuronal hidden states. A constrained inversion of the stochastic model is made possible by assuming power law form for the cross-spectra of the state noise (resp. observation noise) – motivated from previous work on neuronal activity (Beggs & Plenz, 2003; Shin & Kim, 2006; Stam & de Bruin, 2004) – as follows:

$$g_v(\omega,\theta) = \alpha_v \omega^{-\beta_v}$$
$$g_e(\omega,\theta) = \alpha_e \omega^{-\beta_e} \tag{3}$$

Here, $\{\alpha,\beta\} \subset \theta$ are the parameters controlling the amplitudes and exponents of the spectral density of the state noise and the observation noise. The parameterisation of endogenous fluctuations means that the states are no longer probabilistic; hence the inversion scheme is significantly simpler, requiring estimation of only the parameters (and hyperparameters) of the model.

Standard Bayesian model inversion is used to infer the parameters of the model in (1), (2) and (3) from the observed signal $y(t)$. The description of the Bayesian model inversion procedures based on a variational Laplace scheme can be found elsewhere for the interested readers (K. Friston, Mattout, Trujillo-Barreto, Ashburner, & Penny, 2007; K. J. Friston, Harrison, & Penny, 2003; Razi & Friston, 2016).

### Parametric Empirical Bayes

Empirical Bayes refers to the Bayesian inversion or fitting of hierarchical models. In hierarchical models, constraints on the posterior density over model parameters at any given level are provided by the level above. These constraints are called empirical priors because they are informed by empirical data. We recently introduced a second-level or between-subjects model over parameters, which represents how individual (within-subject) connections derive from the subjects' group membership (K. J. Friston et al.,

2016a) – based on parametric empirical Bayes (PEB). This approach calls on Bayesian Model Reduction (BMR) (K. J. Friston et al., 2016b) to finesse the inversion of multiple models of a single dataset or a single (hierarchical) model of multiple datasets. BMR allows one to compute posterior densities over model parameters, under new prior densities, without explicitly inverting the model again. For example, one can invert a DCM for each subject in a group and then evaluate the posterior density over group effects, using the posterior densities over parameters from the single subject inversion. This may improve subject-specific parameter estimates, by using group-level estimates to rescue individual DCM from local optima. Mathematically, for DCM studies with *N* subjects and *M* parameters per DCM, we have a hierarchical model, where the responses of the *i*-th subject and the distribution of the parameters over subjects can be modelled as:

$$y_i = \Gamma_i^{(1)}(\theta^{(1)}) + \varepsilon_i^{(1)} \qquad (4)$$

$$\theta^{(1)} = \Gamma^{(2)}\left(\theta^{(2)}\right) + \varepsilon^{(2)}$$

$$\theta^{(2)} = \eta + \varepsilon^{(3)} ,$$

where, $y_i$ is the BOLD time series from *i-th* subject *and* $\Gamma_i^{(1)}$ is a nonlinear mapping from the parameters of a model to the predicted response $y$ for e.g. as shown in Eq. 1 and Eq. 2 above. $\varepsilon_i^{(1)}$ is independent and identically distributed (i.i.d.) observation noise. In this hierarchical form, *empirical priors* encoding second (between-subject) level effects place constraints on subject-specific parameters. The second level would be a linear model where the random effects are parameterised in terms of their precision:

$$\Gamma^{(2)}\left(\theta^{(2)}\right) = (X \otimes W)\beta ,$$

where $\beta \subset \theta$ are group means or effects encoded by a design matrix with between-subject ($X$) and within-subject ($W$) parts. The between-subject part encodes differences among subjects or covariates such as age, while the within-subject part specifies mixtures of parameters that show random effects. We assume that the first column of the design matrix is a constant term modelling group means and subsequent columns encode group differences or covariates such as age.

### Subjective Effects (5D-ASC)

Two subcategories of oceanic boundlessness (ego dissolution) were measured on the retrospective 5D-ASC 360 minutes after the administration of psilocybin. These subcategories were blissful state and changed meaning of precepts. Under 0.2 mg/kg oral psilocybin, we found group level blissful state, average = 44.23/100; SD = 35.87; range = 3.33–100, changed meaning of precepts average = 37.56/100; SD = 36.32; range = 0.00–100, and anxiety average = 4.28/100; SD = 6.60; range = 0.00–22.00.

Under placebo we found group level blissful state, average = 2.85/100; SD = 4.81; range = 0.00–16, changed meaning of precepts average = 0.45/100; SD = 1.20; range = 0.00–5.33, and anxiety average = 0.69/100; SD = 2.65; range = 0.00–11.83.

### Self-connections in DCM

Please note that in DCM, the self-connections are always modelled as inhibitory (to preclude any run-away excitation), but these parameters in the model are log-scaled for the sake of numerical stability of the model fitting procedures. This (log) scaling means that these self-connections can take both positive (red) and negative values (blue). A positive self-connection means a relative increased inhibition, whereas a negative self-connection means a relative decreased inhibition (i.e., disinhibition). Inhibitory self-connections control the regions' gain or sensitivity to inputs. Decreased self-inhibition suggests increased synaptic gain or sensitivity to inputs, while increased self-inhibition suggests reduced synaptic gain or sensitivity to inputs. Importantly, only the self-connections are log-scaled in DCM. See Supplementary Fig. 2 for region connectivity matrices.

### ROI Identification using Neurosynth

Terms used to identify ROI coordinates using neurosynth automated meta analysis (https://neurosynth.org/analyses/terms/): posterior cingulate, medial prefrontal, anterior cingulate, anterior insula, dlpfc, posterior parietal, amygdala.

MRI Data Acquisition and Preprocessing Additional Details

T1-weigthed images were collected via a 3D magnetization-prepared rapid gradient-echo sequence (MP-RAGE) with the following parameters: voxel size= 0.7x0.7x0.7 mm3, time between two inversion pulses= 3123 ms, inversion time= 1055 ms, inter-echo delay= 12 ms, flip angle= 8°, matrix= 320x335, field of view= 224x235 mm2, 236 sagittal slices. Furthermore T2-weighted images were collected using via a turbo spin-echo sequence with the following parameters: voxel size= 0.7x0.7x0.7 mm3, repetition time= 2500 ms, echo time= 415 ms, flip angle= 90°, matrix= 320x335, field of view= 224x235 mm2, 236 sagittal slices.

**Figure S1.**

$$\begin{bmatrix} 1 & 0 & 0 \\ 1 & 0 & 0 \\ 1 & 1 & 0 \\ 1 & 1 & 0 \\ 1 & 0 & 1 \\ 1 & 0 & 1 \end{bmatrix}$$

**Design matrix used to infer changes from placebo.** Our design matrix designated the placebo group to serve as the baseline. Regressors in our design matrix encode: 1) placebo group 2) the additive effect of being in the second group (psilocybin after 70 min) relative to the placebo group.

**Figure S2.**

**A)** **B)**

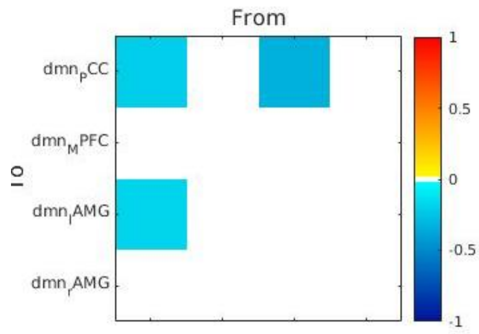


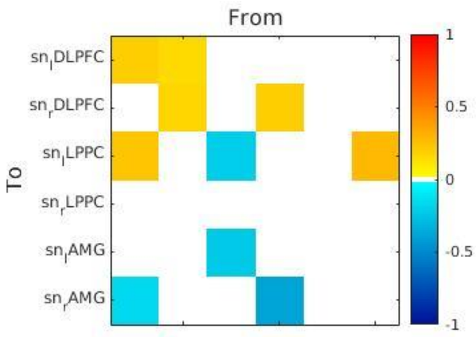


**C)**

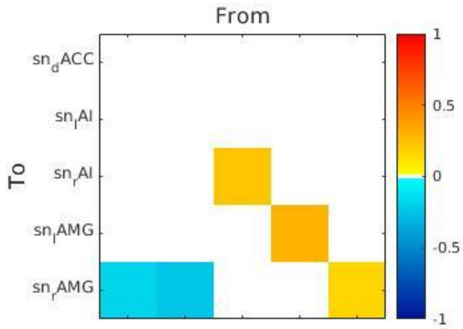


**Group level effective connectivity (EC) change from placebo 70 min post psilocybin administration.**

A) DMN B) CEN C) SN change from placebo EC. Posterior probability threshold = 0.99. See Table S1 for posterior expectations (effect size) and credible intervals.

**Figure S3.**

**A) Blissful state** **Changed meaning of precepts**

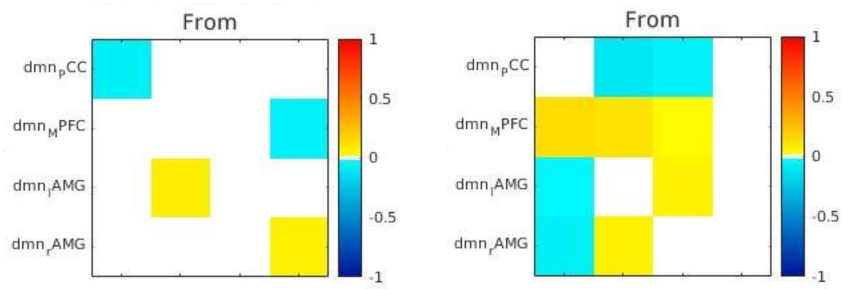


**B)**

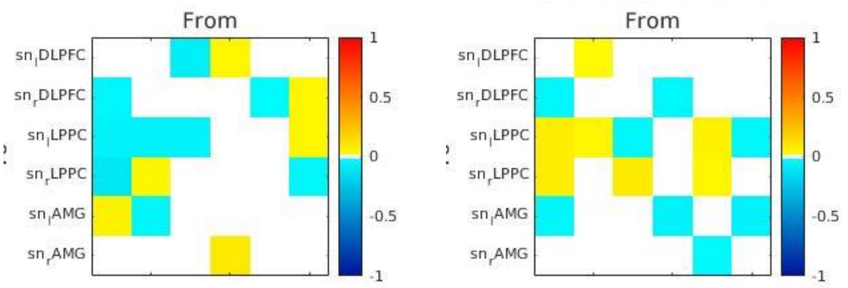


**C)**

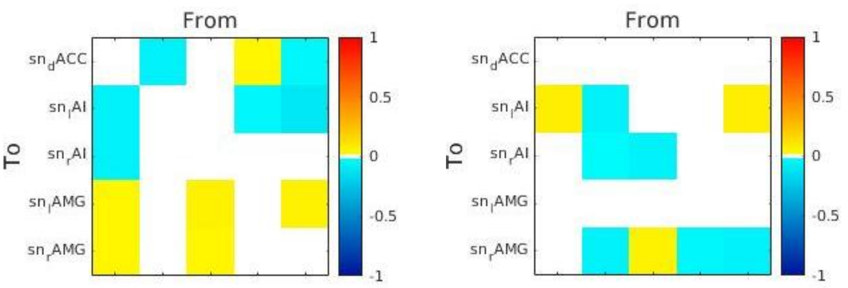


**Behavioural associations to group level effective connectivity (EC) change from placebo 70 min post psilocybin administration.** A) DMN B) CEN C) SN change from placebo EC. Posterior probability threshold = 0.99.

**Table S1.**

| DMN-AMG Connections | Valence & Effect Size | Credible Intervals (low/high) |
|---|---|---|
| PCC → PCC | -0.21 | -0.321/-0.095 |
| PCC → lAMG | -0.18 | -0.270/-0.080 |
| lAMG → PCC | -0.33 | -0.460/-0.201 |
| *CEN-AMG Connections* | *Valence & Effect Size* | *Credible Intervals (low/high)* |
| rLPPC → rDLPFC | 0.20 | 0.118/0.283 |
| rLPPC → rAMG | -0.39 | -0.512/-0.261 |
| lLPPC → lLPPC | 0.20 | -0.384/-0.024 |
| lLPPC → lAMG | -0.22 | -0.344/-0.104 |
| rDLPFC → rDLPFC | 0.17 | 0.040/0.301 |
| rDLPFC → lDLPFC | 0.15 | 0.074/0.232 |
| lDLPFC → lDLPFC | 0.19 | 0.051/0.329 |
| lDLPFC → lLPPC | 0.23 | 0.096/0.363 |
| lDLPFC → rAMG | -0.16 | -0.300/-0.028 |
| rAMG → lLPPC | 0.28 | 0.167/0.384 |
| *SN-AMG Connections* | *Valence & Effect Size* | *Credible Intervals (low/high)* |
| dACC → rAMG | -0.19 | -0.310/-0.063 |
| rAI → rAI | 0.23 | 0.131/0.320 |
| lAI → rAMG | -0.24 | -0.362/-0.117 |
| lAMG → lAMG | 0.31 | 0.153/0.464 |
| rAMG → rAMG | 0.16 | 0.046/0.269 |

**Between regions effective connectivity change from placebo 70 min post psilocybin administration.**

All results are for posterior probability > 0.99.


Beggs, J. M., & Plenz, D. (2003). Neuronal avalanches in neocortical circuits. *J Neurosci, 23*(35), 11167-11177. Retrieved from https://www.ncbi.nlm.nih.gov/pubmed/14657176

Friston, K., Mattout, J., Trujillo-Barreto, N., Ashburner, J., & Penny, W. (2007). Variational free energy and the Laplace approximation. *Neuroimage, 34*(1), 220-234. doi:10.1016/j.neuroimage.2006.08.035

Friston, K. J., Harrison, L., & Penny, W. (2003). Dynamic causal modelling. *NeuroImage, 19*(4), 1273-1302. doi:10.1016/s1053-8119(03)00202-7

Friston, K. J., Kahan, J., Biswal, B., & Razi, A. (2014). A DCM for resting state fMRI. *NeuroImage, 94*, 396-407. doi:https://doi.org/10.1016/j.neuroimage.2013.12.009

Friston, K. J., Litvak, V., Oswal, A., Razi, A., Stephan, K. E., van Wijk, B. C., . . . Zeidman, P. (2016a). Bayesian model reduction and empirical Bayes for group (DCM) studies. *Neuroimage, 128*, 413-431. doi:10.1016/j.neuroimage.2015.11.015

Friston, K. J., Litvak, V., Oswal, A., Razi, A., Stephan, K. E., van Wijk, B. C. M., . . . Zeidman, P. (2016b). Bayesian model reduction and empirical Bayes for group (DCM) studies. *NeuroImage, 128*, 413-431. doi:10.1016/j.neuroimage.2015.11.015

Razi, A., & Friston, K. (2016). The Connected Brain: Causality, models, and intrinsic dynamics. *IEEE Signal Processing Magazine, 33*(3), 14-35.

Shin, C. W., & Kim, S. (2006). Self-organized criticality and scale-free properties in emergent functional neural networks. *Phys Rev E Stat Nonlin Soft Matter Phys, 74*(4 Pt 2), 045101. doi:10.1103/PhysRevE.74.045101

Stam, C. J., & de Bruin, E. A. (2004). Scale-free dynamics of global functional connectivity in the human brain. *Hum Brain Mapp, 22*(2), 97-109. doi:10.1002/hbm.20016
